# Within-Frequency Temporal Processing and Speech Perception in Cochlear Implant Recipients and Normal Hearing Listeners

**DOI:** 10.1101/2020.07.21.20159137

**Authors:** Chelsea M. Blankenship, Jareen Meinzen-Derr, Fawen Zhang

**Author notes:** **Corresponding Author:** Address correspondence to Chelsea Blankenship, Communication Sciences Research Center, Cincinnati Children’s Hospital Medical Center, 3333 Burnet Avenue, Cincinnati, Ohio 45229, USA.

## Abstract

**Objective:** Speech recognition performance among cochlear implant (CI) recipients is highly variable and is influenced by their ability to perceive rapid changes within the acoustic signal (i.e., temporal resolution). A behavioral gap detection test is commonly used to assess temporal processing however it requires active participation, and therefore may be infeasible for young children and individuals with disabilities. Alternatively, cortical auditory evoked potentials (CAEPs) can be elicited by a silent gap embedded in a longer duration stimulus and have been used as an objective measure of temporal resolution. Only a few studies have examined within-frequency gap detection (identical pre- and post-gap frequency), most of which were conducted with normal hearing (NH) individuals and did not include speech perception. The purpose of the study is to evaluate behavioral and electrophysiological measures of within-frequency temporal processing and speech perception in NH and CI recipients.

**Design:** Eleven post-lingually deafened adult CI recipients (n = 15 ears; mean age = 50.4 yrs.) and eleven age- and gender-matched NH individuals (n = 15 ears; mean age = 49.0 yrs.) were recruited. Speech perception was assessed with the CNC word test, AzBio sentence test, and BKB Speech-in-Noise test. Within-frequency (2 kHz pre- and post-gap tone) behavioral gap detection thresholds (GDT) were measured using an adaptive, two-alternative, forced-choice paradigm. Within-frequency CAEPs were measured using four gap duration conditions based on the individual’s behavioral GDT including a supra-threshold (GDTx3), threshold (GDT), sub-threshold (GDT/3), and reference (no gap) condition. Mixed effect models examined group differences in speech perception, behavioral GDTs, and CAEP amplitude and latency. Correlation analyses examined the relationship between the CAEP response, behavioral measures of speech perception and temporal processing, and demographic factors.

**Results:** CI recipients had significantly poorer speech perception scores with no significant differences in behavioral within-frequency GDTs compared to NH participants. CI recipients had poorer CAEP waveform morphology, smaller N1, larger P2 amplitude, and increased P1 latency compared to NH participants. Additionally, older participants displayed smaller N1-P2 amplitude compared to younger participants. Bivariate group correlation analysis showed that individuals with poorer within-frequency GDTs displayed significantly poorer performance on the AzBio sentences in noise and BKB Speech-in-Noise test. Multivariate canonical correlation analysis showed a significant relationship between the within-frequency CAEP amplitude and latency and behavioral measures of speech perception and temporal processing.

**Conclusions:** CI recipients had poorer speech understanding in quiet and noise yet similar behavioral GDTs compared to NH participants. NH participants showed the anticipated trend of increased N1-P2 amplitude as CAEP gap duration increased. However, CAEP amplitude and latency remained relatively stable across gap duration conditions for CI recipients. Instead, significant group and age effects for CAEP peak amplitude and latency were found that can likely be attributed to differences in cortical neuron density, adaptation, and recovery between the groups. Lastly correlation analysis indicates that individuals with poorer temporal processing are likely to have adequate speech perception in quiet but worse speech understanding in noise.

## INTRODUCTION

Accurate auditory perception is essential for many processes such as environmental awareness, speech perception, music appreciation, and localization. It requires precise temporal resolution throughout the peripheral and central auditory nervous system, starting with initial processing at the neuronal level to more complex functions such as speech perception and language processing (Mauk and Buonomano 2004; Shinn et al. 2009). With regard to speech perception, the ability to precisely segregate acoustic information is critical for accurate perception of phonemes which then build to form syllables, words, and sentences (Mauk and Buonomano 2004). Individuals with cochlear implants (CIs) often display poorer temporal processing skills compared to normal hearing (NH) individuals (Blankenship et al. 2016; Muchnik et al. 1994; Tyler et al. 1989). Given the importance of temporal resolution in auditory perception and overall speech understanding, it is reasonable to anticipate that deficits in temporal processing will be associated with poorer speech and language outcomes after implantation.

Temporal resolution refers to the ability to perceive rapid changes within the acoustic signal over time (Moore 2012). Speech is a complex acoustic signal that is comprised of many temporal cues which the auditory system must be able to encode, resolve, and integrate in a meaningful manner to allow accurate speech perception (Mauk and Buonomano 2004). For example, stop consonants contain acoustic content that vary rapidly over time and are distinguished based on place and manner of articulation (Tallal et al. 1993). A defining feature of stop consonants is the voice onset time, which is the time between the initial consonantal burst to the onset of voicing. The voice onset time can be short as in voiced stop consonants (5 to 25 ms) or around 50 to 105 ms as in unvoiced (Lisker and Abramson 1964; Phillips 1999). The ability of the auditory system to detect and integrate the voice onset time with other acoustic features is essential for listeners to differentiate similar phonemes, consonants, and syllables such as /da/ vs. /ta/ or /ba/ vs. /pa/ (Holt and Lotto 2010; Mauk and Buonomano 2004). Furthermore, the ability to perceive the duration of frequency transitions (/ba/ vs /wa/) and the silent period between consonants and vowels (/sa/ vs /sta/) are fundamental for accurate phoneme discrimination (Dorman et al. 1979; Liberman et al. 1956). Subsequently accurate identification of phonemes is necessary for comprehension of syllables that occur every 200 to 400 ms in continuous speech (Mauk and Buonomano 2004) which is essential for overall speech understanding. At a more global level, voiced speech contains periodic glottal pulses, from which the temporal spacing is used by the auditory system to identify voice pitch or the fundamental frequency of the speaker. This information is used to help differentiate the age and gender of the speaker, where males tend to have a fundamental frequency ranging from 85 to 155 Hz and females from 165 to 255 Hz. Subsequent changes in the rate of the fundamental frequency/glottal pulse rate are employed when making statements (rapidly declining intonation contours), asking questions (rising contours), conveying emotion or attitude, and are the basis for melodic contours (Phillips 1999). Thus, a deficit in temporal processing can prohibit accurate speech understanding.

Temporal resolution is traditionally examined using a behavioral psychoacoustic gap detection task. In this paradigm, the listener is presented with two stimuli, one of which is the target and contains a brief silent period or “gap” and the other is continuous (“no gap”). The participant is instructed to listen to both sounds and indicate which contains a silent gap. An adaptive procedure is used to measure the shortest gap of silence the listener can detect, which is deemed their gap detection threshold (GDT). When the silent gap is long enough, the listener is easily able to parse the sound into three segments: the pre-gap marker, gap of silence, and the post-gap marker. However, as the silent gap duration nears threshold, the task becomes more difficult and the gap is perceived as a “glitch” or “hiccup,” or is inaudible (Moore et al. 1993). GDTs often vary substantially among studies due to the sheer number of factors including stimulus level, marker bandwidth and center frequency, spectral similarity of markers, and presentation mode that can affect results (Eddins et al. 1992; Lister et al. 2007; Lister and Roberts 2005; Plomp 1964; Schneider and Hamstra 1999). The lowest GDT thresholds can be obtained when the stimuli are presented at supra-threshold intensity level (Eddins et al. 1992; Moore et al. 1993), using sinusoidal stimuli (Moore and Glasberg 1988) in the within-frequency (spectrally identical pre- and post-gap markers) condition (Lister et al. 2002; Lister and Roberts 2005). Across-frequency gap detection (spectrally dissimilar pre- and post-gap markers) has been proposed to be more representative of the speech due to the inclusion of both frequency and temporal changes. However, across-frequency gap detection is harder and results in larger and more variable GDTs. Since across-frequency temporal processing is not the main focus of this manuscript, only within-frequency studies will be discussed further.

In NH listeners, within-frequency GDTs have been reported as low as 2-3 ms (Blankenship et al. 2016; Eddins et al. 1992; Heinrich and Schneider 2006; Phillips et al. 1998; Plomp 1964). In contrast, CI recipients display a wide range of GDTs with some individuals performing comparable to their NH peers (Bierer et al. 2015; Shannon 1989) and others displaying much larger GDTs (Blankenship et al. 2016; Tyler et al. 1989; Wei et al. 2007). Tyler et al. (1989) reported GDTs in CI recipients that ranged from 7.5 to 200 ms and noted that CI recipients with smaller GDTs (< 40 ms) displayed better speech recognition abilities and environmental awareness than individuals with larger GDTs (> 40 ms). Similarly, Muchnik et al. (1994) categorized CI recipients into a low GDT (< 35 ms) and high GDT (> 35 ms) group and reported that CI recipients with open-set speech recognition had lower GDTs. Blankenship et al. (2016) reported GDTs that ranged from 2 to 100 ms in CI recipients which were significantly correlated with speech perception performance (CNC Phoneme, AzBio in Quiet, BKB-SIN SNR-50). Although there is no clinical definition of good and poor GDTs, it is widely accepted that better temporal processing skills are associated with smaller GDTs, which are required for good speech perception.

Behavioral GDTs are often easily obtained in adults, however, additional factors such as memory, cognition, motivation, and attention affect the feasibility of behavioral testing procedures in difficult-to-test adults and children. In addition, approximately 30 to 40% of pediatric CI recipients have disabilities or medical conditions that prohibit their ability to provide reliable behavioral responses (Chilosi et al. 2010). Thus, an objective electroencephalographic (EEG) measure that does not rely on a behavioral response, would be extremely beneficial for difficult-to-test populations. EEG assessment is a safe and non-invasive measure that has been used extensively to assess the function of the human auditory system (Harris et al. 2012; Picton 2011, 2013). It can measure millisecond-by millisecond neural activity and ultimately provide precise information to monitor and diagnose temporal processing deficits. Therefore, electrophysiological assessment can be used as a non-behavioral measure to provide information about how the auditory system processes temporal information.

In recent years, cortical auditory evoked potentials (CAEPs) have been used to study the neural detection of temporal changes in the acoustic stimulus in individuals with normal hearing, elderly adults, auditory neuropathy, and cochlear implants (Atcherson et al. 2009; Harris et al. 2012; He et al. 2012; He et al. 2013; He et al. 2015; Lister et al. 2007; Lister et al. 2011; Michalewski et al. 2005; Palmer and Musiek 2013, 2014; Pratt et al. 2005; Pratt et al. 2007). Most studies examined within-frequency electrophysiological GDT using broadband, narrowband, or Guassian noise stimuli. In NH adults, the CAEP response has been recorded to gap durations as small as 2 ms (Palmer and Musiek 2013) but are more commonly present for gap durations ≥ 5 ms (Atcherson et al. 2009; He et al. 2012; Michalewski et al. 2005; Pratt et al. 2005). With regard to stimulus frequency, poorer GDTs have been reported for 0.5 kHz narrowband noise stimuli than those observed for 1 and 4 kHz (Atcherson et al. 2009). Compared to normal hearing listeners, individuals with known temporal processing deficits, such as auditory neuropathy, display CAEP responses that were only present for longer gap durations ranging from 10 to 100 ms (He et al. 2015; Michalewski et al. 2005). In addition, older adults display significantly poorer behavioral and electrophysiological GDTs with larger variability compared to younger adults (Harris et al. 2012; Lister et al. 2011; Palmer and Musiek 2014). In CI recipients, the only electrophysiological GDT study was completed in children with auditory neuropathy using direct electrical stimulation (800 ms biphasic pulses) instead of acoustic stimuli. While some participants displayed electrophysiological GDTs between 5 to 10 ms, several individuals had much higher GDTs (Range = 20 to 100 ms).

The effect of gap duration on the amplitude and latency of the CAEP response has been well studied. CAEP response amplitude increases as a function of the saliency of the gap duration (Atcherson et al. 2009; He et al. 2012; Lister et al. 2007; Michalewski et al. 2005; Palmer and Musiek 2013; Pratt et al. 2005) and gap durations that are clearly audible (supra-threshold) generate repeatable CAEP responses while inaudible gap durations (sub-threshold) generally do not evoke a clear CAEP response (Lister et al. 2007). However, the effect of gap duration on CAEP latency is not as clear. Pratt et al. (2005) and Lister et al. (2007) reported that the CAEP peak latency decreased as the gap duration increased, however, other studies have reported no effect (Michalewski et al. 2005; Palmer and Musiek 2013). He et al. (2012) reported a non-monotonic effect where CAEP latency decreased for gap durations up to 20 ms but increased for longer gap durations. Due to the consistent effect of increased CAEP response amplitude with larger gap durations, CAEP amplitude is a more reliable indicator of auditory discrimination and potential objective indicator of temporal processing at the level of the auditory cortex.

A close association between electrophysiological and behavioral GDTs has been reported for both NH adults (Atcherson et al. 2009; He et al. 2012; Palmer and Musiek 2014; Pratt et al. 2005) and individuals with auditory neuropathy (He et al. 2015; Michalewski et al. 2005). In young NH adults, Atcherson et al. (2009) reported a mean difference of 6.6 ms at 1 kHz and 2.8 ms at 4 kHz between electrophysiological and behavioral GDTs. Palmer and Musiek (2014) reported an even smaller mean difference of 0.4 ms between electrophysiological and behavioral GDTs in both younger and older adults. Michalewski et al. (2005) reported that while most individuals with auditory neuropathy display comparable behavioral and electrophysiological GDTs, there were 5 participants for which an electrophysiological GDT was not able to be obtained (> 50 ms) despite a measurable behavioral GDT (Range = 5 to 40 ms). In a pediatric CI candidate cohort (n = 15), He et al. (2015) also reported that one participant with auditory neuropathy had a behavioral GDT of 10 ms with a much larger electrophysiological GDT of 100 ms. Therefore, it might be the case that some individuals with temporal processing impairments display larger discrepancies between behavioral and electrophysiological GDTs.

Another important consideration is the relationship between speech perception and electrophysiological GDTs, which few studies have addressed. In adults with auditory neuropathy, Michalewski et al. (2005) reported the general trend that individuals with poorer temporal processing (larger electrophysiological GDTs) had poorer speech perception scores, however, no statistical analysis was completed. In a group of pediatric CI candidates (He et al. 2013) and CI recipients (He et al. 2015) with auditory neuropathy, a robust negative correlation was reported between open-set word recognition scores and electrophysiological GDTs (ρ = −0.81, *p* < 0.01). Individuals with poorer word recognition scores required larger gap durations (> 20 ms) to elicit a CAEP response.

In summary, electrophysiological GDTs show great potential for use as an objective measure to evaluate neural processing of temporal changes within an acoustic stimulus. Several studies have demonstrated a consistent effect of gap duration on CAEP response amplitude and have shown high correlations between behavioral and electrophysiological GDT. However, limited research has been conducted with individuals with known temporal processing deficits, such as CI recipients. Additionally, only a few studies have included measures of speech perception. Given the importance of temporal resolution to auditory perception and speech understanding and the aforementioned gaps in the literature, the present study was undertaken: (1) evaluate within-frequency temporal processing using the CAEP response evoked by silent gaps of various durations; (2) examine within-frequency behavioral GDTs and common measures of speech perception; and (3) explore the correlation among behavioral GDTs, speech perception, demographic characteristics, and CAEP amplitude and latency.

## MATERIALS AND METHODS

### Participants

Eleven adult CI recipients (Mean = 50.4 yrs; Range = 25.2 to 68.3) and eleven age- and gender-matched NH adults (Mean = 49.0 yrs; Range = 24.7 to 68.5) were enrolled in the study. The age difference between the NH and CI participants ranged from 0 to 4.3 years (Mean = 1.6 rs). Four CI recipients were bilaterally implanted and each ear was tested separately, for a total of 15 CI ears examined in the study. The NH participants were tested in the same ear as their age- and gender-matched CI user. Therefore, four NH participants were tested in both their left and right ear separately and seven NH individuals were tested in only one ear, for a total of 15 NH ears included in the study. All CI recipients were post-lingually deafened (onset of bilateral severe-to-profound hearing loss > 3 years of age), implanted with Cochlear Americas Implant System (Cochlear Americas Ltd, New South Wales, Australia) and had a minimum of one year experience with their CI to allow optimization of speech processor settings (Holden et al. 2013). CI participants reported full time use of their CI during all waking hours. Relevant CI recipient demographic and device information is displayed in Table 1. All NH and CI participants were right-handed (Edinburgh Handedness Inventory; Oldfield 1971), native speakers of American English, and did not report a history of neurological or psychiatric disorders, or brain injury. The research study was approved by the Institutional Review Board of the University of Cincinnati. Informed consent was obtained from all participants and they were paid for participation.

**Table 1.**
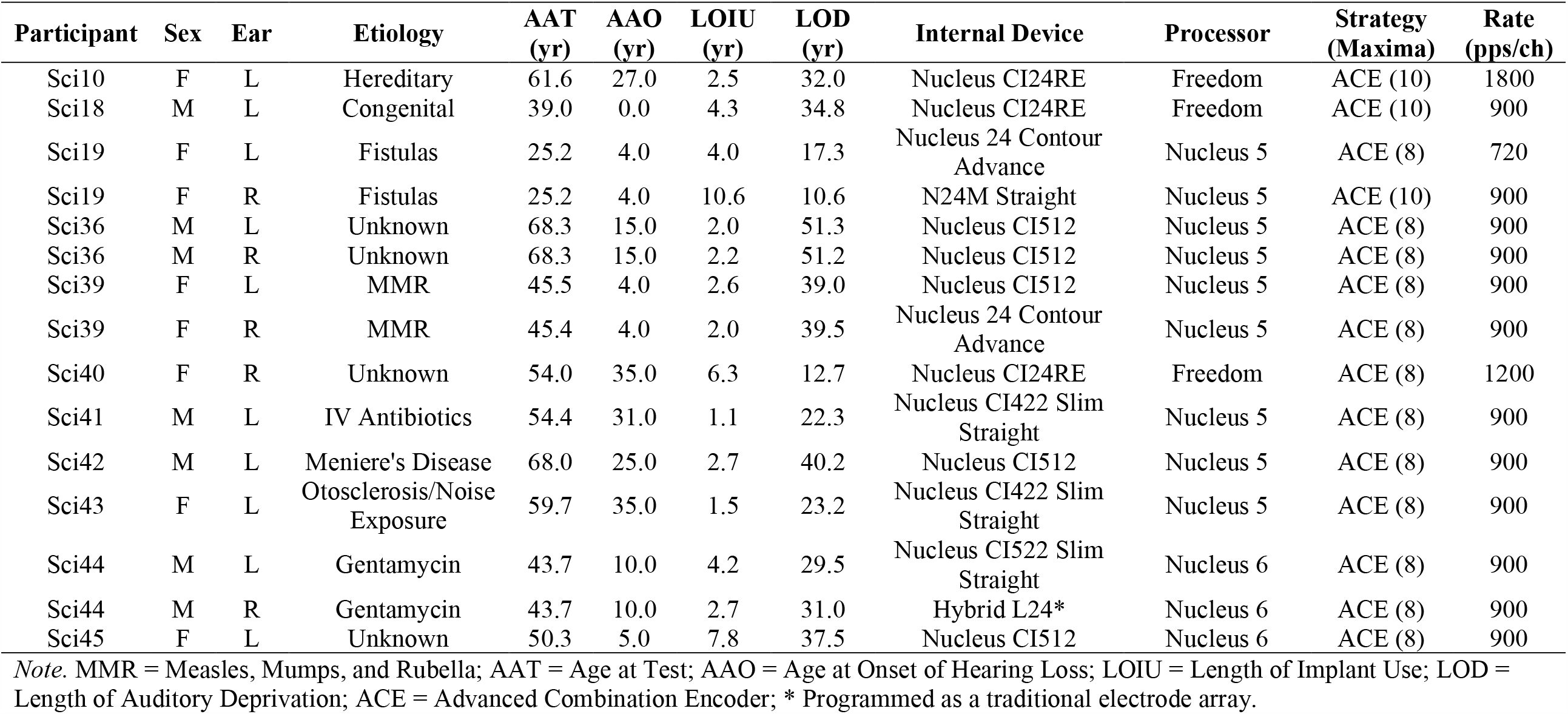
Cochlear Implant Recipient Demographic and Device Information.

### Stimuli

The within-frequency stimuli used for behavioral and EEG testing were 2 kHz pure-tones created using Audacity software (version 1.2.5; opensource, http://audacity.sourceforge.ent) with

a sampling rate of 44.1 kHz. Pure-tone stimuli were used instead of the more common narrow or broadband noise stimuli, as studies have shown that pure-tone GDTs are less affected by stimulus complexity (Heinrich et al. 2014; Heinrich and Schneider 2006) and result in a more accurate measure of temporal resolution (Moore and Glasberg 1988). The 2 kHz pure-tone markers varied in duration from 250 to 350 ms to prevent duration cues to aid in behavioral gap detection (Formby and Forrest 1991; Lister et al. 2007). The stimuli included a 10 ms rise-time for the pre- and 10 ms fall-time for the post-gap marker. A silent gap ranging in duration from 0 to 120 ms was inserted in between the pre- and post-gap markers (0 ms indicates no gap, 2-20 ms in 2 ms increments, 20-120 in 5 ms increments) and included a 1 ms rise- and fall-time around the silent gap (offset of pre-gap marker, onset of post-gap marker). These stimuli and rise-fall times are similar to those used in previous studies (Lister et al. 2007; Lister and Roberts 2005; Phillips et al. 1997). For EEG testing, the within-frequency stimuli were identical to that used for the behavioral testing, except gap durations ranged from 0 to 360 ms in 1 ms increments and the marker duration was fixed at 300 ms for both the pre- and post-gap marker. EEG testing requires a fixed stimulus duration to allow averaging of the time-locked neural responses across stimulus presentations. A longer duration marker (300 ms) was used to prevent overlapping neural responses to the responses evoked by the onset of the pre- and post-gap marker.

### General Study Procedures

Testing was completed in the Human Auditory Evoked Potential Lab at the University of Cincinnati over the course of one or two sessions depending on participant’s preference and if one ear (3 hours of testing) or both ears were tested (6 hours of testing). All participants completed behavioral testing first followed by EEG testing within a double-walled sound treated booth (Industrial Acoustics Company, North Aurora, Illinois) that meets the standard for acceptable room noise for audiometric rooms (ANSI 1999). CI recipients completed all testing with their speech processors turned on and adjusted to their everyday settings (volume, sensitivity, and program) which were held constant throughout the entire test session(s). For NH individuals and unilaterally implanted CI recipients, testing was completed with the contralateral ear occluded with an E-A-R disposable foam ear plug to prevent participation. For bilaterally implanted recipients, testing was completed with the contralateral speech processor removed. All testing was completed at the Most Comfortable Level (MCL; 7 on a 0-10 loudness scale; Hoppe et al., 2001). Testing at the MCL is commonly used in CI research since it allows easier comparison of test results between NH and CI participants in the current and future studies (Bierer et al. 2015; Dorman et al. 2014; Han et al. 2005). The presentation order of the test materials, and the lists used within each test, was randomized to minimize order effects.

### Hearing Threshold Assessment

All NH participants completed otoscopy to ensure an unobstructed ear canal and a healthy appearing tympanic membrane. Tympanometry was completed using a GSI TympStar Version 2 (Grason Stradler, Eden Prairie, MN) with a 226 Hz probe tone. All NH participants displayed type A tympanograms (Compliance = 0.3 to 1.7 ml, Gradient = 50 to 110 daPa, Tympanometric Peak Pressure = −150 to +100 daPa) indicative of normal middle ear status (Jerger 1970; Margolis and Hunter 1999). Next audiometric thresholds were measured (GSI-61 or AudioStar Pro; Grason Stradler, Eden Prairie, MN) to ensure a 25-30 dB SL necessary for optimal gap detection (Fitzgibbons and Gordon-Salant 1987) when stimuli were presented at individual MCL. For NH participants, thresholds were measured at octave test frequencies from 0.25 to 8 kHz with pulsed pure-tones and the Hughson-Westlake procedure with a 5 dB step size (Carhart and Jerger 1959) using insert ER-3A (Etymotic Research, Elk Grove Village, IL) earphones. For CI recipients, audiometric thresholds were measured using frequency-modulated tones from 0.25 to 6 kHz due to standing waves with soundfield presentation at 8 kHz.

### Speech Perception Assessment

Speech perception performance was measured using the Minimum Speech Test Battery for Adult Cochlear Implant Users (CNC, AzBio, BKB-SIN), which was designed for the clinical and research assessment of open-set word and sentence recognition in quiet and in noise (Etymotic 2005; Peterson and Lehiste 1962; Spahr et al. 2012). The Consonant-Nucleus-Consonant Word Test (CNC) assesses open-set monosyllabic word recognition in quiet. Two CNC word lists per test ear were administered to each participant. Responses were scored based on the entire word (% correct; CNC-Word) and number of phonemes (% correct; CNC-Phoneme) repeated correctly (Peterson and Lehiste 1962). The Arizona Biomedical Sentence Recognition Test (AzBio) assesses open-set sentence recognition in quiet and noise. The sentences are spoken in a conversational speaking style by two male and two female talkers providing limited contextual cues with each list equated for intelligibility. Participants were presented with two lists in quiet and two lists in noise (+10 dB signal-to-noise ratio [SNR]) per test ear (Spahr et al. 2012). Responses were scored based on the percentage of words repeated correctly for sentences in quiet (AzBio-Quiet) and noise (AzBio-Noise). The Bamford-Kowal-Bench Speech-in-Noise Test (BKB-SIN) is an adaptive measure of open-set sentence recognition with SNRs that ranged from +21 to −6 dB. Two BKB-SIN word list pairs were administered to each participant per test ear (Etymotic 2005). The purpose of the BKB-SIN is to determine the SNR in decibels necessary for the individual to understand 50% of the sentence (SNR-50). Acoustic stimuli were presented monaurally through insert earphones to NH listeners and through a loudspeaker placed at 0 degrees azimuth 1 meter from the CI recipients. Participants were instructed to verbally repeat what they heard and did not receive feedback based on their responses.

### Temporal Processing Assessment

A behavioral gap detection task was used to assess within-frequency temporal processing abilities. Stimuli were presented using APEX software (Francart et al. 2008) with an adaptive, two-alternative forced-choice procedure with an up-down stepping rule. For each trial, the participant was presented with two sounds, one that contained a silent gap (target) and the other without a gap (reference). The presentation order of the target and reference stimuli was randomized and the duration between presentations was set at 0.5 seconds. For each trial, the participant was instructed to select the sound that contained the silent gap. Visual feedback was provided, and testing continued until five reversals were completed and the mean of the last three reversals was recorded as the GDT.

### Electrophysiological Recording

EEG stimuli were presented using Neuroscan STIM^2^ software and recordings were collected with a Neuroscan™ recording system (SCAN software version 4.3, Compumedics Neuroscan, Inc., Charlotte, NC) paired with a NuAmps digital amplifier. Continuous EEG activity was recorded with a sampling rate of 1000 Hz using a 40-channel Neuroscan Quik-Cap (Compumedics Neuroscan, Inc., Charlotte, NC) organized according to the extended 10-20 International system. The reference electrode was placed on the earlobe contralateral to the test ear, which has been found to reduce stimulus artifact in some CI recipients (Liang et al. 2017; McNeill et al. 2007). Electrooculography was recorded using four electrodes placed 1 cm above and below the left eye (vertical) and at the outer canthus of each eye (horizontal). For CI recipients, approximately one to three electrodes surrounding the transmission coil were not used during recording. Electrode impedances were kept below 5 kΩ and were balanced across electrodes. EEG recordings were collected for a total of four gap duration conditions including: (1) threshold (behavioral GDT), (2) supra-threshold (behavioral GDT x 3), (3) sub-threshold (behavioral GDT/3), and (4) reference (no gap). Calculated gap durations were rounded to the nearest whole integer. The order of stimulus conditions was randomized within and across participants. Continuous EEG recordings were collected with a minimum of 200 and 400 stimulus trials collected from NH and CI recipients, respectively. The inter-stimulus interval was fixed at 0.9 seconds and triggers were time-locked to the onset of the pre-gap marker. For all participants, the EEG stimuli were presented at MCL (7 on a 0-10 loudness scale, Hoppe et al., 2001) through a sound-field speaker placed 1 meter from the test ear at 90 and −90 degrees azimuth corresponding to the right and left ear respectively. During EEG testing, participants were seated

in a comfortable chair, instructed to ignore the stimuli, and relax but stay awake during the EEG test session.

### Data Analysis

#### Behavioral Data

The CNC word (CNC-Word and CNC-Phoneme) and AzBio sentence (AzBio-Quiet, AzBio-Noise) speech perception tests were evaluated in terms of percent correct. For the BKB-SIN, the SNR-50 was calculated which determines the SNR necessary to understand 50% of the key words contained in the sentence. The within-frequency GDT was calculated as the mean gap duration (ms) of the last three reversals in the psychoacoustic gap detection task.

#### Electrophysiological Data

Initial offline analysis was completed within Neuroscan software version 4.3 with subsequent analysis completed in EEGLAB 13.6.5b (http://sccn.ucsd.edu/eeglab) run under Matlab R2017b (The Mathworks, Natick, MA). In the Neuroscan 4.3 analysis software, the EEG data was digitally band-pass filtered (0.1 to 30 Hz with a 6 dB/octave roll-off), epoched from −100 to 200 ms beyond the offset of the post-gap marker, and baseline corrected using the pre-stimulus window (−100 to 0 ms). Note the epoch length varied among participants for the sub-threshold, threshold, and supra-threshold conditions because EEG stimuli were based on individual behavioral GDT thresholds. In EEGLAB, any bad or unused electrodes surrounding the speech processor coil were removed and data epochs were visually analyzed to remove approximately 10% of the epochs that contained non-stereotyped artifact such as body movement. Independent Component Analysis (ICA) was used to decompose the EEG data into mutually independent components including those from neural and artifactual sources. Independent components that represented artifacts arising from ocular movement, electrode, or cochlear implant artifact were identified and removed through the visual inspection of component properties including the waveform, 2-D voltage map, and the spectrum (Delorme and Makeig 2004; Delorme et al. 2007). Deleted channels were interpolated, electrodes were re-referenced to the average reference (Delorme and Makeig 2004; Hagemann et al. 2001), baseline corrected using the pre-stimulus window (−100 to 0 ms), and filtered from 0.1 to 30 Hz using a band-pass Fast Fourier Transform linear filter.

Visual inspection of the scalp topography verified that the CAEP response was most easily identified along the central/midline electrodes (Fz, FC3, FCz, FC4, Cz). To aide in peak identification for CI recipients, data from the aforementioned five electrodes were averaged together to form one final averaged EEG waveform (Harris et al. 2012; Michalewski et al. 2005). An averaged waveform was derived in response to each gap duration condition resulting in a total of four waveforms per participant. For each averaged waveform, CAEP peak components (P1, N1, and P2) were identified for both the pre- and post-gap markers. For the pre-gap marker, the P1 was identified as the maximum positive peak between 25 and 75 ms, N1 was the maximum negative peak between 75 and 150 ms, and P2 was the maximum positive peak occurring between 150 and 220 ms that was morphologically appropriate (Harris et al. 2012; Martin 2007). For the post-gap marker conditions, the response latency might vary depending on the saliency of the silent gap duration (i.e., sub-threshold or threshold conditions would have increased latency compared to the pre-gap P1-N1-P2), and participant group (NH vs CI). Therefore, individual pre-gap marker conditions were used as a guide when identifying post-gap P1-N1-P2 amplitude and latency. The presence of P1, N1, and P2 was determined by visual inspection of the waveforms by two reviewers (Blankenship and Zhang) on which they agreed for all waveforms. The P1, N1, P2 amplitude were measured as the change in amplitude from baseline to maximum negative or positive peak. The N1-P2 amplitude was calculated as the change in amplitude measured from N1 trough to the P2 peak. For CAEP responses with a broad peak or trough, the midpoint was chosen to measure the response amplitude and latency.

#### Statistical Analysis

Descriptive statistics were calculated for all outcome variables with measures of central tendency and dispersion for interval variables. Boxplots were created to study the distribution of the outcomes and identify outliers (values > 1.5 x interquartile range). For behavioral measures, multiple mixed effect models were used to examine differences in speech perception (CNC-Word, CNC-Phoneme, AzBio-Quiet, AzBio-Noise, SNR-50) and within-frequency behavioral GDTs between the NH and CI recipients. Participant group (NH and CI) and test ear (left and right) were included as fixed effects, age at test was included as a covariate, and participant ID was included as a random effect to account for participants that were tested in both ears separately. For post-gap CAEP amplitude and latency, multiple mixed effect models were conducted with participant group (NH and CI), gap duration condition (sub-threshold, threshold, supra-threshold), and test ear (left and right) included as fixed effects with age at test as a covariate, and participant ID as a random effect. The Satterthwaite approximation method was used to estimate degrees of freedom and the Holm’s step-down adjustment method was applied for pairwise comparisons. Models were compared with and without outliers and if the main findings changed, a description was included in the results section.

Multivariate canonical correlations were conducted to assess relationships among post-gap CAEP (P1 and N1-P2 amplitude and P1, N1, P2 latency), Behavioral (CNC-Word, CNC-Phoneme, AzBio-Quiet, AzBio-Noise, SNR-50, Within-GDT), and Demographic variables (age at test, age at onset of HL, length of implant use, length of auditory deprivation). Canonical correlations that included CAEP variables resulted in separate analyses for each gap duration condition (sub-threshold, threshold, and supra-threshold). Therefore a total of seven separate canonical correlation analyses were completed with the following pairs: Behavioral and Demographic (CI data only), Demographic and CAEP (CI data only; sub-threshold, threshold, supra-threshold), Behavioral and CAEP (NH and CI data; sub-threshold, threshold, supra-threshold). Bivariate scatter plots were examined to evaluate the assumptions of linearity, multivariate normality, and homoscedasticity. One-tailed spearman rank correlations coefficients were used to assess the strength of pairwise correlations between within-frequency GDTs and speech perception (CNC, AzBio, BKB-SIN). Data were analyzed using SPSS statistical software (IBM Corp. Released 2019. IBM SPSS Statistics for Windows, Version 26.0. Armonk, NY: IBM Corp.).

## RESULTS

### Within-Frequency Behavioral Results

Group mean audiometric thresholds for NH and CI participants are displayed in Figure 1. NH individuals had thresholds that were ≤ 25 dB from 0.25 to 8 kHz, per the NH inclusion criteria. CI recipients had thresholds that ranged from 15 to 45 dB HL across all test frequencies, within the clinical normal to moderate hearing loss range. The purpose of obtaining audiometric thresholds in both the NH and CI group was to ensure a 25 to 30 dB SL recommended for optimal gap detection (Fitzgibbons and Gordon-Salant 1987). The presentation level ranged from 25 to 45 dB SL for the CI group (Mean = 37 dB) and ranged from 43 to 55 dB SL (Mean = 48 dB) for the NH group.

**Figure 1.**
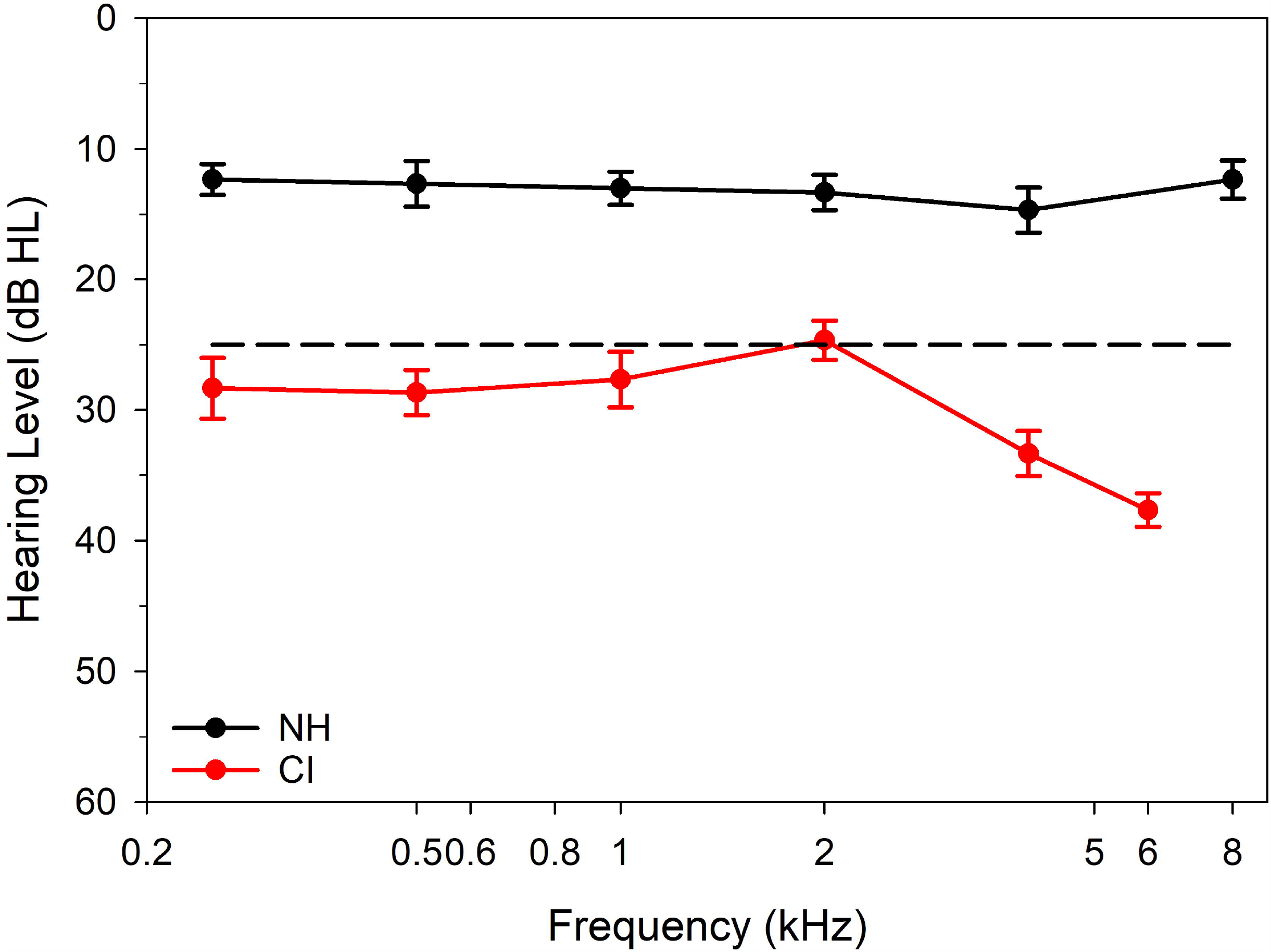
Mean audiometric thresholds for NH (n = 15 ears) and CI recipients (n = 15 ears) at octave test frequencies from 0.25 to 8 kHz. The black dashed line indicates normal hearing criteria and error bars represent 95% confidence intervals (ANSI 2010).

Group mean speech perception scores and within-frequency gap detections thresholds are displayed in Table 2. Mixed effect model analysis results are displayed in Table 3. NH participants performed at ceiling level (individual scores ≥ 97 %) on all non-adaptive speech perception tests. CI recipients showed variable performance across speech perception measures with only one CI recipient that performed near ceiling level (≥ 95%) for the CNC-Phoneme score and all CNC-Word scores ≤ 85%. Four CI recipients (5 ears) performed at ceiling level (≥ 95%) on the AzBio-Quiet but for the AzBio-Noise, only one individual scored close to ceiling level at 94.3% while all other participants scored < 86%. On the adaptive BKB-SIN test, the mean SNR-50 (e.g., SNR at which they could understand 50% of the target words) for NH participants was - 0.7 dB, while the CI recipients required a much higher SNR-50 of +8.2 dB. On the within-frequency gap detection task, all NH participants had a GDT of 2 ms which was the smallest gap duration on the task. Most CI participants also had a behavioral GDT of 2 ms, except for two CI recipients (one bilaterally implanted) for a total of three CI ears (Sci36-Left = 51.7 ms, Sci36-Right = 26.7 ms, Sci43-Left = 41.7 ms).

**Table 2.** Within-Frequency Behavioral Gap Detection and Speech Perception Performance.

| **Behavioral Measures** | **NH Group** |  |  | **CI Group** |  |  |
| --- | --- | --- | --- | --- | --- | --- |
|  | **Mean ± SD** | **Minimum** | **Maximum** | **Mean ± SD** | **Minimum** | **Maximum** |
| CNC-Phoneme (%) | 99.7 ± 0.4 | 98.7 | 100.0 | 84.8 ± 9.3 | 62.7 | 95.0 |
| CNC-Word (%) | 99.1 ± 1.0 | 97.0 | 100.0 | 70.5 ± 12.8 | 43.0 | 85.0 |
| AzBio-Quiet (%) | 99.5 ± 0.6 | 98.3 | 100.0 | 88.5 ± 8.9 | 73.9 | 99.6 |
| AzBio-Noise* (%) | 99.2 ± 0.8 | 97.5 | 100.0 | 65.5 ± 19.2 | 38.1 | 94.3 |
| SNR-50 (dB) | -0.7 ± 1.2 | -3.0 | 1.8 | 8.2 ± 3.7 | 2.8 | 13.3 |
| Within-GDT (ms) | 2.0 ± 0.0 | 2.0 | 2.0 | 9.6 ± 16.4 | 2.0 | 51.7 |

*Note.* SNR-50 = Signal-to-Noise Ratio required for 50% correct; Within-GDT = Within-Frequency Gap Detection Threshold; \* AzBio-Noise was only completed for 14 CI ears due to equipment issues. All other tests include data from 15 NH and 15 CI ears.

**Table 3.** Behavioral and Across-Frequency Post-Gap CAEP Mixed Effect Model Analysis (*p-*values, F-test, degrees of freedom displayed).

|  | Group | Condition | Test Ear | Age at Test |
| --- | --- | --- | --- | --- |
| **Behavioral Measures** |  |  |  |  |
| CNC-Phoneme | < ***0.001*** | — | 0.359 | 0.779 |
| F (DF = 5 to 12) | 26.955 | — | 1.012 | 0.083 |
| CNC-Word | < ***0.001*** | — | 0.531 | 0.902 |
| F (DF = 3 to 8) | 54.280 | — | 0.483 | 0.017 |
| AzBio-Quiet | < ***0.001*** | — | 0.850 | 0.749 |
| F (DF = 8 to 12) | 20.008 | — | 0.037 | 0.110 |
| AzBio-Noise | < ***0.001*** | — | 0.279 | 0.280 |
| F (DF = 11 to 15) | 42.645 | — | 1.260 | 1.288 |
| SNR-50 | < ***0.001*** | — | 0.438 | 0.116 |
| F (DF = 14 to 17) | 72.136 | — | 0.630 | 2.816 |
| Within-GDT | 0.158 | — | 0.381 | 0.152 |
| F (DF = 10 to 19) | 2.157 | — | 0.837 | 2.239 |
| **CAEP Measures** |  |  |  |  |
| P1 Amplitude | 0.217 | 0.804 | 0.593 | 0.088 |
| F (DF = 13 to 69) | 1.653 | 0.218 | 0.288 | 3.406 |
| N1 Amplitude | ***0.011*** | 0.176 | 0.766 | 0.097 |
| F (DF = 17 to 73) | 7.932 | 1.792 | 0.089 | 3.074 |
| P2 Amplitude | ***0.018*** | 0.829 | 0.238 | 0.640 |
| F (DF = 13 to 66) | 6.978 | 0.188 | 1.418 | 0.229 |
| N1-P2 Amplitude | 0.154 | 0.199 | 0.181 | ***0.041*** |
| F (DF = 19 to 74) | 2.187 | 1.662 | 1.823 | 4.802 |
| P1 Latency | ***0.009*** | 0.760 | 0.948 | 0.673 |
| F (DF = 17 to 64) | 8.718 | 0.276 | 0.004 | 0.184 |
| N1 Latency | 0.160 | 0.530 | 0.088 | 0.876 |
| F (DF = 18 to 66) | 2.152 | 0.642 | 3.001 | 0.025 |
| P2 Latency | 0.655 | 0.132 | 0.175 | 0.810 |
| F (DF = 18 to 65) | 0.206 | 2.098 | 1.881 | 0.059 |

*Note.* SNR-50 = Signal-to-Noise Ratio required for 50% correct; Within-GDT = Within-Frequency Gap Detection Threshold; Bold italics indicate significant *p*-values.

Mixed effect model results showed CI participants scored significantly poorer (*p* < 0.001) on all speech perception measures (CNC-Phoneme, CNC-Word, AzBio-Quiet, AzBio-Noise, and SNR-50) compared to NH individuals but a corresponding group difference in within-frequency GDT (*p* = 0.158) was not observed. Fixed effects of test ear and age at test did not reach significance in any model (*p* > 0.05). Although descriptive statistics identified several outliers for behavioral measures of temporal processing and speech perception, mixed effect models with outliers excluded resulted in no change in the main effect significance levels.

### Within-Frequency Electrophysiological Results

ICA is routinely used in the literature to remove artifact from EEG recordings (Delorme et al. 2007; Zhang et al. 2013). Figure 2 displays a CAEP waveform from one NH and one CI participant before and after ICA, displaying the successful removal of CI artifact. As shown in the figure, there are two-time frames of interest, one after the onset of the pre-gap marker and the second after the onset of the post-gap marker. ICA was able to successfully remove CI artifact from all CI participant data.

**Figure 2.**
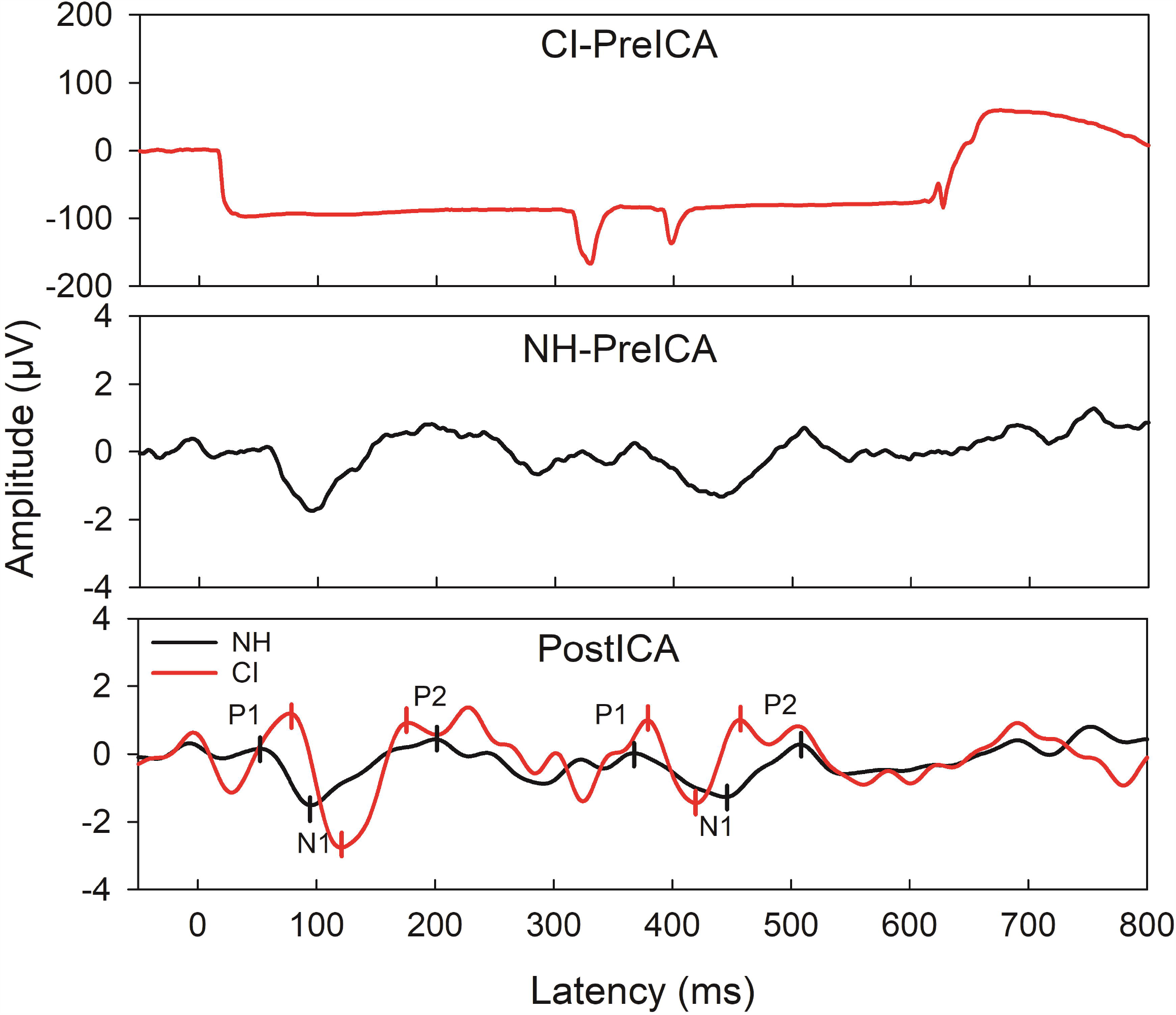
Pre- and post-ICA CAEP waveforms from one CI user (Sci45) and their age- and gender-matched NH control for the within-frequency supra-threshold condition (6 ms gap). The top two figures display the CAEP before ICA for the CI and NH participants respectively and the bottom figure displays the NH and CI waveform after ICA. After ICA, the pre- and post-gap CAEP are apparent for both the NH and CI user. The pre- and post-gap P1-N1-P2 are marked for both the NH and CI participant in the bottom figure.

For the pre-gap marker, the CAEP response was readily identified for all NH and CI participants for all conditions. Since the pre-gap stimuli are identical across conditions (i.e., 2 kHz pure-tone), waveforms were averaged across conditions resulting in one waveform per group and are displayed in Figure 3. Compared to the NH group, CI recipients displayed smaller amplitude and increased latency for P1, N1, and P2 components.

**Figure 3.**
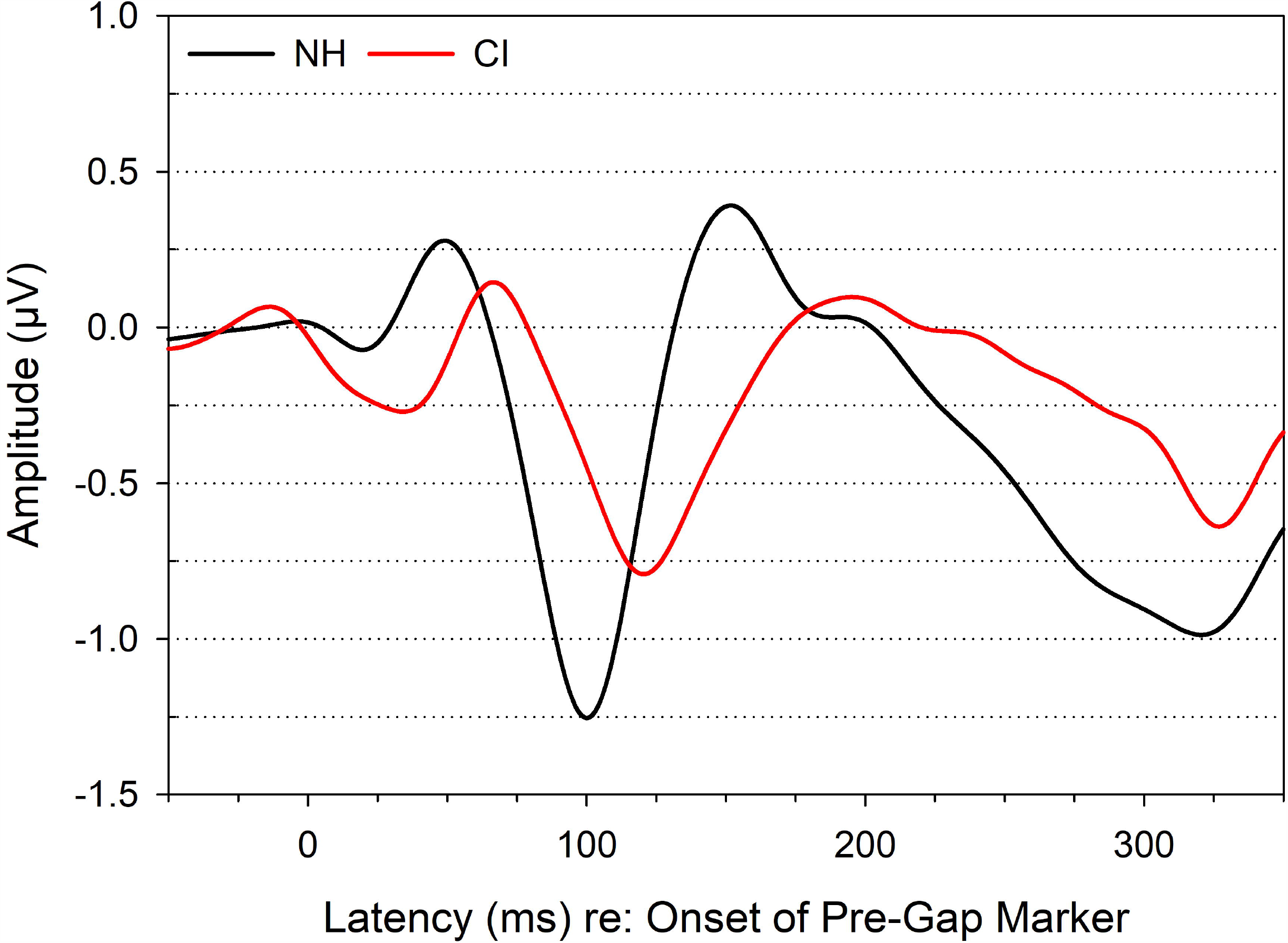
NH and CI group mean pre-gap CAEP waveforms for all within-frequency conditions (reference, sub-threshold, threshold, supra-threshold) averaged together.

Post-gap CAEP waveforms for each group and condition are displayed in Figure 4. For NH participants the P1, N1, P2 latency remained stable across conditions, however, the N1-P2 amplitude increased with longer gap durations. CI recipients displayed poorer CAEP morphology and smaller amplitudes across all conditions compared to the NH participants. NH and CI post-gap CAEP mean and standard deviation amplitude and latency values and the number of present CAEPs for each condition are displayed in Table 4.

**Table 4.** Within-Frequency NH and CI Group Mean CAEP Amplitude and Latency Values.

| Marker | Group | Condition | Count<br>(n) | Amplitude (μV)* | | | | Latency (ms)* | | |
| --- | --- | --- | --- | --- | --- | --- | --- | --- | --- | --- |
|  |  |  |  | P1 | N1 | P2 | N1-P2 | P1 | N1 | P2 |
| Pre-Gap | NH | Reference | 15 | $0.4 \pm 0.4$ | $-1.5 \pm 0.9$ | $0.7 \pm 0.4$ | $2.3 \pm 1.4$ | $56.2 \pm 11.8$ | $100.7 \pm 7.2$ | $166.6 \pm 24.7$ |
| | | Sub-threshold | 15 | $0.5 \pm 0.4$ | $-1.3 \pm 1.0$ | $0.8 \pm 0.6$ | $2.0 \pm 1.4$ | $56.1 \pm 16.3$ | $100.9 \pm 8.9$ | $160.5 \pm 22.6$ |
| | | Threshold | 15 | $0.4 \pm 0.6$ | $-1.4 \pm 1.0$ | $0.6 \pm 0.6$ | $2.0 \pm 1.1$ | $56.3 \pm 11.7$ | $99.5 \pm 7.5$ | $154.9 \pm 22.3$ |
| | | Supra-threshold | 15 | $0.6 \pm 0.5$ | $-1.3 \pm 0.6$ | $0.9 \pm 0.9$ | $2.1 \pm 1.2$ | $54.9 \pm 14.5$ | $99.9 \pm 5.6$ | $159.1 \pm 23.9$ |
| | CI | Reference | 15 | $0.3 \pm 0.7$ | $-1.4 \pm 1.7$ | $0.4 \pm 0.8$ | $1.8 \pm 1.3$ | $71.0 \pm 15.6$ | $121.3 \pm 15.3$ | $188.5 \pm 27.5$ |
| | | Sub-threshold | 15 | $0.6 \pm 0.7$ | $-1.0 \pm 0.9$ | $0.6 \pm 0.5$ | $1.6 \pm 0.9$ | $68.7 \pm 20.9$ | $116.7 \pm 27.6$ | $183.0 \pm 24.6$ |
| | | Threshold | 15 | $0.5 \pm 0.6$ | $-0.8 \pm 0.8$ | $0.5 \pm 0.6$ | $1.2 \pm 0.7$ | $70.0 \pm 13.5$ | $123.3 \pm 18.7$ | $192.3 \pm 43.9$ |
| | | Supra-threshold | 15 | $0.1 \pm 1.4$ | $-1.3 \pm 1.6$ | $0.3 \pm 0.7$ | $1.6 \pm 1.4$ | $74.2 \pm 19.5$ | $120.8 \pm 21.2$ | $187.4 \pm 36.6$ |
| Post-Gap | NH | Reference | — | — | — | — | — | — | — | — |
| | | Sub-threshold | 13 | $-0.4 \pm 0.2$ | $-1.6 \pm 0.7$ | $-0.4 \pm 0.5$ | $1.2 \pm 0.7$ | $58.9 \pm 14.8$ | $126.1 \pm 17.9$ | $185.4 \pm 14.4$ |
| | | Threshold | 13 | $-0.4 \pm 0.4$ | $-1.9 \pm 0.8$ | $-0.4 \pm 0.5$ | $1.5 \pm 0.6$ | $56.8 \pm 11.4$ | $120.1 \pm 12.8$ | $192.7 \pm 12.7$ |
| | | Supra-threshold | 15 | $-0.4 \pm 0.6$ | $-2.2 \pm 1.2$ | $-0.3 \pm 0.4$ | $1.8 \pm 1.1$ | $54.5 \pm 10.6$ | $120.7 \pm 12.2$ | $187.1 \pm 15.4$ |
|  | CI | Reference | — | — | — | — | — | — | — | — |
| | | Sub-threshold | 13 | $-0.1 \pm 0.9$ | $-1.1 \pm 0.9$ | $0.0 \pm 0.7$ | $1.2 \pm 1.0$ | $89.2 \pm 38.8$ | $136.2 \pm 41.4$ | $190.7 \pm 42.3$ |
| | | Threshold | 13 | $0.1 \pm 0.9$ | $-0.8 \pm 1.1$ | $0.2 \pm 1.1$ | $1.0 \pm 1.1$ | $93.7 \pm 37.8$ | $146.4 \pm 43.1$ | $207.8 \pm 57.8$ |
| | | Supra-threshold | 14 | $-0.3 \pm 1.2$ | $-1.4 \pm 1.4$ | $-0.2 \pm 0.8$ | $1.2 \pm .9$ | $106.7 \pm 43.1$ | $157.5 \pm 46.8$ | $206.8 \pm 47.0$ |

\*Mean values ± one standard deviation.

**Figure 4.**
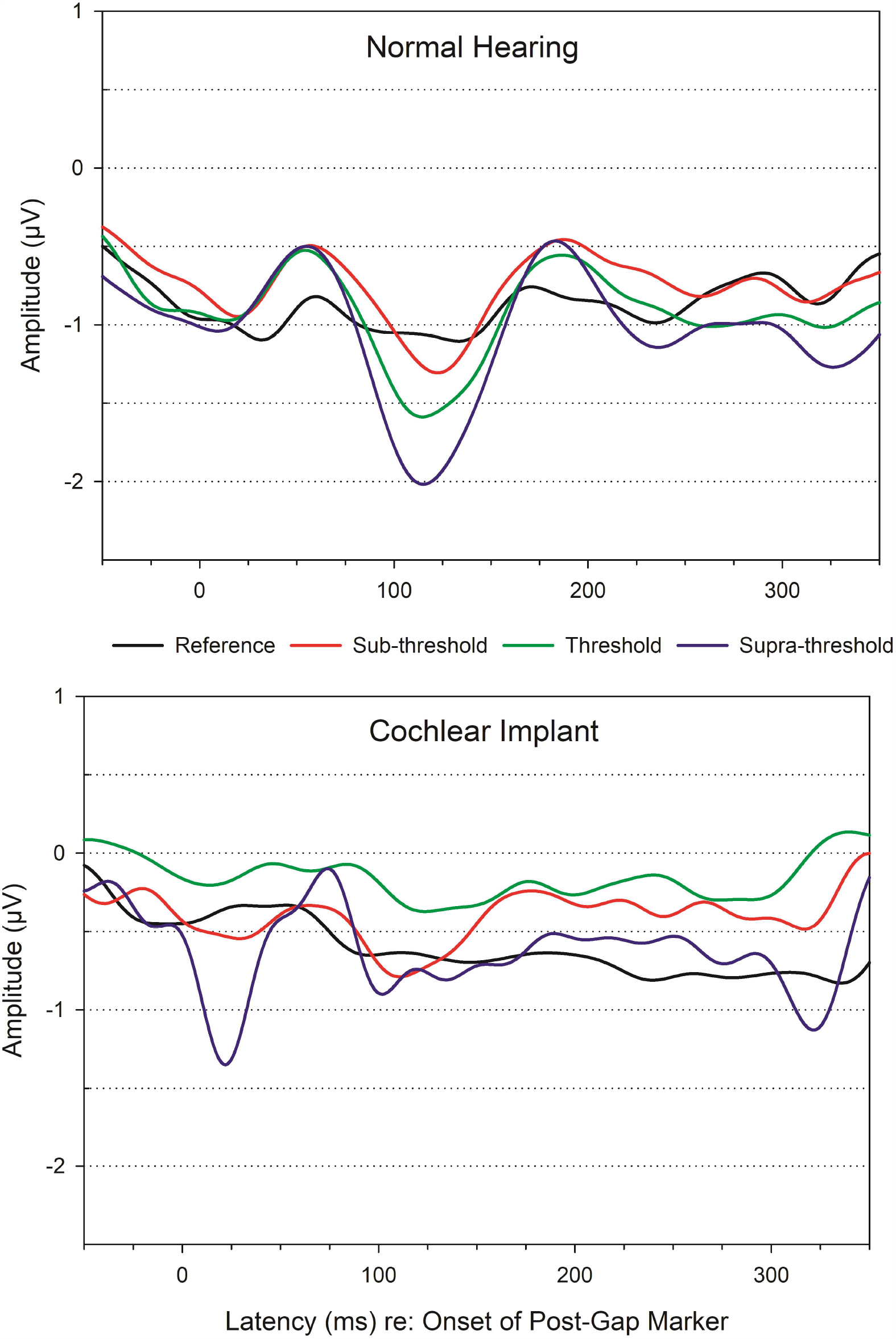
NH and CI group mean post-gap CAEP waveforms displayed for each gap duration condition. The top panel displays waveforms from NH participants and the bottom displays CI participant waveforms.

Multiple mixed effect models were used to evaluate the effect of group (NH and CI), gap duration condition (sub-threshold, threshold, supra-threshold), test ear (left and right), and age at test on CAEP amplitude and latency values. Individual CAEP model significance values are displayed in Table 3. For P1 amplitude, a significant effect of group, condition, ear, or age at test was not observed (*p* > 0.05). A significant group effect was found for N1 amplitude (*p* = 0.011) where NH participants had a significantly larger amplitude (Mean = −1.863 µV) compared to CI recipients (Mean = −1.036 µV). For P2 amplitude, NH participants had a significantly smaller amplitude (Mean = −0.407 µV) compared to CI participants (Mean = 0.034 µV; *p* = 0.018). However, for the N1-P2 peak-to-peak amplitude, which is a more stable measure than the baseline-to-peak amplitude of N1 and P2 (Prosser et al. 1981), a significant group effect was not observed. Instead, a significant effect of age at test indicated that older individuals displayed smaller N1-P2 amplitudes (Mean = 1.183 µV; *p* = 0.041) than younger adults (Mean = 1.505 µV).

For P1 latency, there was a main effect of group (*p* = 0.009), where CI recipients displayed longer latencies (Mean = 90.3 ms) compared to NH participants (Mean = 56.4 ms), however, there was no main effect of gap duration condition, test ear, or age at test. For N1 and P2 latency, a significant effect of group, gap duration condition, test ear, or age at test was not observed (*p* > 0.05).

Descriptive statistics identified several outliers for CAEP amplitude and latency measurements, therefore mixed effect models were rerun with outliers excluded to evaluate their influence on the main effects. There was no change in the significance levels for amplitude (P1, N1, P1, and N1-P2) or P1 and N1 latency. For P2 latency, an effect test ear was significant (F_[1,53.6]_ = 4.1, *p* = 0.047) with the left ear displaying slightly longer latencies (Mean = 192.8 ms) than the right ear (Mean = 182.3 ms).

### Within-Frequency Canonical Correlation Analysis

Multivariate canonical correlations were conducted to assess relationships among CAEP amplitude and latency (sub-threshold, threshold, supra-threshold conditions), behavioral (CNC-Word, CNC-Phoneme, AzBio-Quiet, AzBio-Noise, SNR-50, Within-GDT), and demographic variables (age at test, age at onset, length of implant use, length of auditory deprivation). No significant relationships were found between demographic and CAEP variables or demographic and behavioral variables. For the behavioral and CAEP canonical correlation analysis, both the threshold and supra-threshold CAEP conditions were significant.

The behavioral and CAEP threshold condition correlation analysis supported a two-dimensional relationship. With all five canonical correlations included, Wilks’ lambda was 0.019 (F _[30, 54]_ = 3.0, *p* < 0.001) and with the first correlation removed, Wilks’ lambda was 0.140 (F_[20, 47]_ = 1.9, *p* = 0.034). The first canonical correlation was 0.93 (86% overlapping variance) with an eigen value of 6.19; the second canonical correlation was 0.87 (76% overlapping variance) with an eigen value of 3.20. The CAEP threshold coefficients and cross-loadings for the first and second canonical correlation are displayed in Table 5. The canonical coefficients represent the individual items’ relative contribution to the variate (Tatham et al. 1998). For example, within the first canonical correlation, the order of contribution to the first behavioral variate from largest to smallest is AzBio-Noise, AzBio-Quiet, CNC-Phoneme, Within-GDT, CNC-Word, and SNR-50. For the first CAEP variate, the order of contribution is P2 latency, N1 latency, N1-P2 amplitude, P1 latency, and P1 amplitude. Next, cross-loadings, which provide a direct measure of the behavioral-CAEP relationship, were examined (Dillon and M. 1984; Tatham et al. 1998). For behavioral variables, Within-GDT had the highest correlation of −0.48 indicating that 23% of the variance in Within-GDT is explained by the first CAEP variate. All other speech perception measures displayed much lower correlations, with < 2% of the variance in individual speech perception measures explained by the first CAEP variate. For CAEP variables, N1-P2 amplitude and P1 latency displayed the highest correlations of −0.75 and 0.51 respectively indicating that approximately 56% of variance in N1-P1 amplitude and 26% of the variance in P1 latency is explained by the first behavioral variate. N1 and P2 latency each account for < 10% of the variance and P1 amplitude only accounts for 1% of the variance in behavioral measures.

**Table 5.** Within-Frequency Behavioral and Post-Gap Threshold CAEP Canonical Correlation Analysis.

| Item Content | First Canonical<br>Correlation |  | Second Canonical<br>Correlation |  |
| --- | --- | --- | --- | --- |
|  | Cross-Loading | Coefficient | Cross-Loading | Coefficient |
| **Behavioral** |  |  |  |  |
| CNC-Word | -0.13 | 1.08 | -0.53 | -4.42 |
| CNC-Phoneme | -0.11 | -1.47 | -0.43 | 4.16 |
| AzBio-Quiet | 0.05 | 2.53 | -0.53 | -2.04 |
| AzBio-Noise | -0.08 | -2.54 | -0.48 | 3.33 |
| SNR-50 | 0.14 | 0.27 | 0.52 | 1.62 |
| Within-GDT | -0.48 | -1.14 | 0.06 | -0.02 |
| **CAEP** |  |  |  |  |
| P1 Amplitude | -0.10 | -0.20 | 0.62 | 0.63 |
| N1-P2 Amplitude | -0.75 | -0.46 | 0.01 | 0.15 |
| P1 Latency | 0.51 | 0.36 | 0.55 | 0.35 |
| N1 Latency | 0.26 | 0.56 | 0.67 | 0.24 |
| P2 Latency | -0.30 | -0.76 | 0.58 | 0.23 |

*Note.* SNR-50 = Signal-to-Noise Ratio required for 50% correct; Within-GDT = Within-Frequency Gap Detection Threshold.

Examination of the second canonical coefficients indicate that speech perception measures (CNC, AzBio, SNR-50) all have high levels of contribution with minimal contribution from Within-GDT. For the CAEP variate, P1 amplitude contributes the most followed by P1, N1, P2 latency and N1-P2 amplitude. The behavioral cross-loadings indicate approximately 27 to 28% of the variance in CNC-Word, AzBio-Quiet, SNR-50, 23% of the variance in AzBio-Noise, and 18% of the variance in CNC-Phoneme can be explained by the second CAEP variate. Within-GDT has a minimal contribution with < 1% of the variance in Within-GDT explained by the CAEP variate. The percent of variance in each of the CAEP variables explained by the second behavioral variate is as follows: P1 amplitude (38%), N1-P2 amplitude (<1%), N1 latency (44%), P2 latency (33%), and P1 latency (30%).

Examination of the loadings for the first and second canonical correlation collectively, indicate that the first canonical correlation involves a primary relationship between Within-GDT and N1-P2 amplitude (smaller GDTs result in smaller N1-P2 amplitude). Whereas the second canonical correlation seems to capture an inverse relationship between behavioral measures of speech perception and P1 amplitude, P1, N1, and P2 latency (poorer speech perception result in larger P1 amplitude and increased P1, N1, and P2 latency).

A significant relationship was also found for the behavioral and CAEP supra-threshold condition with canonical correlation analysis supporting a two-dimensional relationship. With all five canonical correlations included, Wilks’ lambda was 0.070 (F_[30, 70]_ = 2.205, *p* = 0.003) and with the first correlation removed, Wilks’ lambda was 0.189 (F_[20, 60]_ = 1.9, *p* = 0.022). The first two canonical correlation coefficients were 0.79 (63% overlapping variance) and 0.72 (52% overlapping variance) with eigenvalues of 1.708 and 1.086, respectively. The CAEP threshold coefficients and cross-loadings for the first and second canonical correlation are displayed in Table 6. For the first canonical correlation, CNC-Word, CNC-Phoneme, and SNR-50 contributed the most to the behavioral variate with minimal contributions from AzBio (Quiet and Noise) and Within-GDT. For the first CAEP variate, N1 latency contributed the most with P1 and N1-P2 amplitude, P1 and P2 latency contributing to a lesser degree. Examination of the cross-loadings showed SNR-50 had the highest correlation of −0.44 indicating that 19% of the variance in SNR-50 is explained by the first CAEP variate. All other behavioral measures had lower correlations resulting in approximately 5 to 10% of the variance in AzBio (Quiet and Noise), only 3% of the variance in CNC scores, and < 1% of the variance in Within-GDT explained by the first CAEP variate. For CAEP variables, P1 amplitude and N1 latency displayed the highest correlations of - 0.60 and −0.54 respectively indicating that approximately 36% of variance in P1 amplitude and 29% of the variance in P1 latency is explained by the first behavioral variate. N1-P2 amplitude and P1 and P2 latency had lower correlations resulting in only 9 to 12% of the variance explained by the first behavioral variate.

**Table 6.** Within-Frequency Behavioral and Post-Gap Supra-threshold CAEP Canonical Correlation Analysis.

| Item Content | First Canonical<br>Correlation |  | Second Canonical<br>Correlation |  |
| --- | --- | --- | --- | --- |
|  | Cross-Loading | Coefficient | Cross-Loading | Coefficient |
| **Behavioral** |  |  |  |  |
| CNC-Word | 0.18 | -4.17 | 0.08 | 1.69 |
| CNC-Phoneme | 0.17 | 2.37 | 0.05 | -0.58 |
| AzBio-Quiet | 0.24 | 0.10 | 0.01 | -0.95 |
| AzBio-Noise | 0.32 | -0.34 | -0.04 | 1.10 |
| SNR-50 | -0.44 | -2.82 | 0.06 | 0.81 |
| Within-GDT | 0.02 | 0.59 | 0.60 | 1.04 |
| **CAEP** |  |  |  |  |
| P1 Amplitude | -0.60 | -0.66 | 0.20 | 0.65 |
| N1-P2 Amplitude | 0.31 | -0.13 | 0.13 | 0.17 |
| P1 Latency | -0.34 | 0.53 | -0.16 | 0.67 |
| N1 Latency | -0.54 | -1.28 | -0.15 | -2.06 |
| P2 Latency | -0.35 | 0.19 | 0.24 | 1.58 |

*Note.* SNR-50 = Signal-to-Noise Ratio required for 50% correct; Within-GDT = Within-Frequency Gap Detection Threshold.

For the second canonical correlation behavioral variate, the order of contribution from largest to smallest is CNC-Word, AzBio-Noise, Within-GDT, AzBio-Quiet, CNC-Phoneme, SNR-50, and CNC-Phoneme. For the second CAEP variate, N1 and P2 latency contributed the most with P1 and N1-P2 amplitude contributing to a lesser degree. Cross-loadings indicated that Within-GDT had the highest correlation of −0.60 indicating that 36% of the variance in gap detection is explained by the second CAEP variate. All other behavioral measures had much smaller correlation values resulting in < 1% of variance in CNC (Word and Phoneme), AzBio (Quiet and Noise) and SNR-50 scores explained by the second CAEP variate. All CAEP variables showed weak correlations, with each CAEP variable explaining < 5 % of the variance the second Behavioral variate.

Interpretation of the first and second canonical correlation loadings together indicate that the first canonical correlation involves a relationship between speech understanding in noise and CAEP variables and the second canonical correlation involves a relationship between gap detection and CAEP variables. Specifically, better speech understanding in noise is related to decreased P1 amplitude, increased N1-P2 amplitude, and decreased P1, N1, P2 latency. For the second variate, larger gap detection thresholds are related to increased P1 and N1-P2 amplitude, decreased P1 and N1 latency and increased P2 latency.

### Within-Frequency Bivariate Correlation Analysis

Non-parametric spearman rank correlation coefficients were used to assess the strength of pairwise correlations between within-frequency behavioral GDT and speech perception measures (CNC, AzBio, BKB-SIN) for the NH and CI group collectively. Within-frequency GDT and speech perception scatter plots are displayed in Figure 5. A Bonferroni correction was applied to account for multiple comparisons (n = 5), and therefore *p* ≤ 0.01 was considered statistically significant. Pairwise correlations revealed a significant negative correlation for AzBio sentences in Noise (ρ = −0.43, *p* = 0.010) and a significant positive correlation for SNR-50 (ρ = 0.43, *p* = 0.008) with within-frequency GDTs. However, the significant correlation appears to be driven by three data points, which will be reviewed further in the discussion (Sci36-Left =51.7 ms, Sci36-Right = 26.7 ms, Sci43-Left = 41.7 ms). In contrast, correlations between within-frequency GDT and CNC-Phoneme (ρ = −0.31, *p* = 0.047), CNC-Word (ρ = −0.33, *p* = 0.039), and AzBio-Quiet (ρ = −0.38, *p* = 0.020) were not significant (*p* > 0.01).

**Figure 5.**
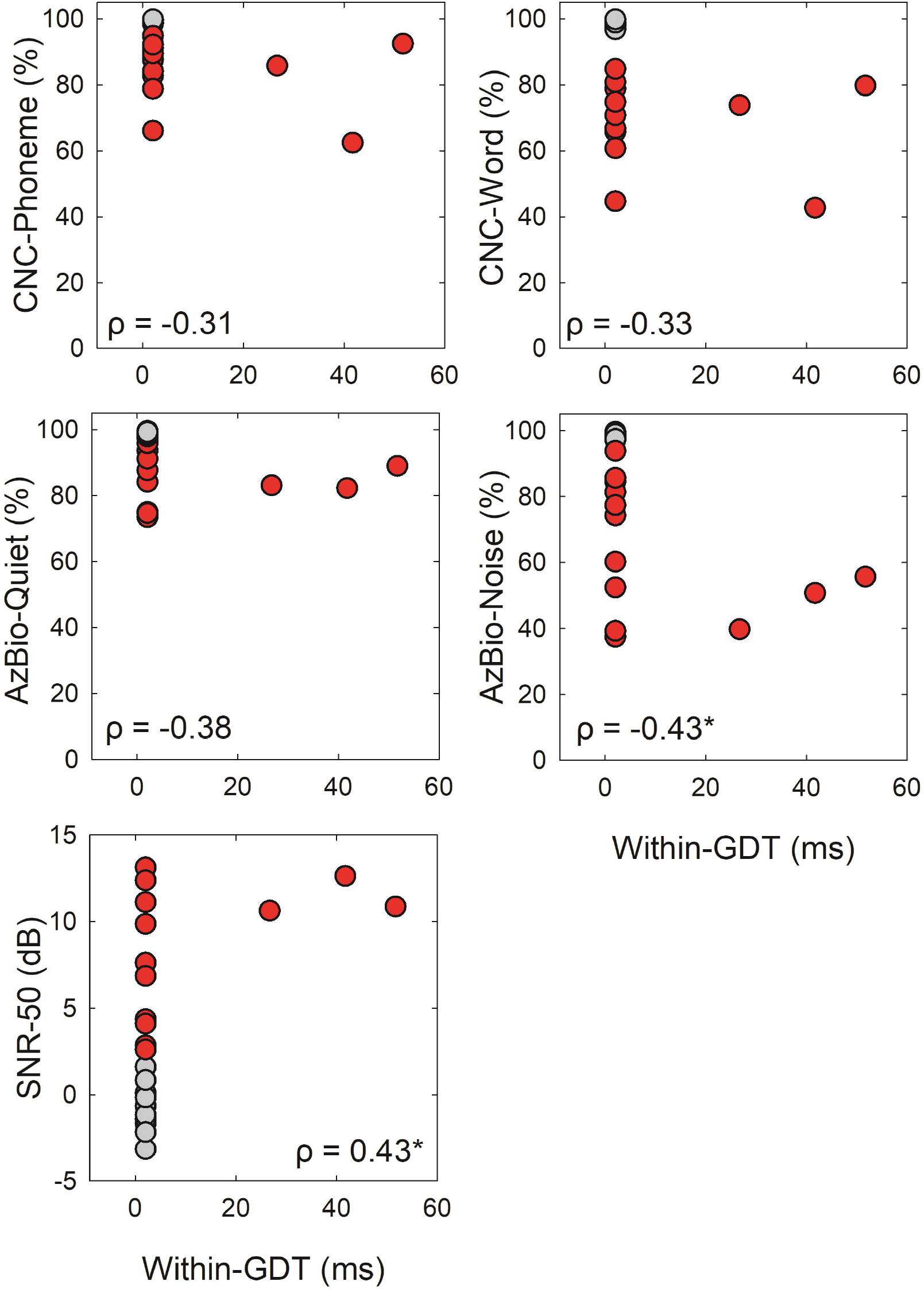
Speech perception scores plotted as a function of within-frequency behavioral GDTs. NH participants are displayed in grey and CI recipients in red. Results of a spearman’s ranked correlation test are displayed in each graph with significant comparison after a Bonferroni correction marked with an asterisk (*p* ≤ 0.01).

## DISCUSSION

The purpose of the present study was to examine behavioral and electrophysiological within-frequency temporal processing and speech perception in CI recipients and individuals with NH. CI recipients had significantly poorer speech perception scores with no significant differences in behavioral within-frequency GDTs compared to NH participants. CAEP results displayed a significant effect of age at test for N1-P2 amplitude and group effect for N1 amplitude, P2 amplitude, and P1 latency. No significant differences in CAEP amplitude and latency were found for gap duration condition (sub-threshold, threshold, supra-threshold) or test ear. Canonical correlations showed the highest correlation occurring between the CAEP threshold condition and behavioral measures of speech perception and temporal processing. Bivariate group correlation analysis showed a significant correlation between within-frequency GDTs and speech perception in noise (AzBio-Noise and BKB-SIN).

### Within-Frequency Behavioral Gap Detection

NH participants performed at ceiling level on the within-frequency gap detection task with all participants displaying a GDT of 2 ms. Our results are similar to Heinrich and Schneider (2006) that reported 1 and 2 kHz pure-tone within-frequency GDTs that ranged from 0.9 to 1.6 ms for younger and 1.2 to 2.5 ms for older participants. Blankenship et al. (2016) reported slightly higher 2 kHz pure-tone within-frequency GDTs that ranged from 2 to 15 ms for young NH listeners with a mean of 5.8 ms. In contrast, Lister et al. (2007) reported within-frequency GDTs that ranged from 7 to 15 ms (Mean = 9.8 ms, SD = 0.8) in young NH adults using 2 kHz narrowband noise. More recently, Alhaidary and Tanniru (2019) reported mean within-frequency GDTs (2 kHz narrowband noise) in NH adults (Mean = 3.8 ms, SD = 2.6 ms) that were comparable to previous studies but slightly higher than the GDTs obtained in NH adults in the current study. Previous studies have shown that GDTs increase with stimulus complexity and shorter marker durations which could explain the slightly higher GDTs reported by previous studies (Alhaidary and Tanniru 2019; Blankenship et al. 2016; Lister et al. 2007).

CI recipients displayed a slightly higher but non-significant group mean within-frequency GDTs of 9.6 ms (SD = 16.4) that ranged from 2 to 51.7 ms. Our results are consistent with the acoustic GDTs reported in the literature for post-lingually deafened CI users (Blankenship et al. 2016; Muchnik et al. 1994; Tyler et al. 1989; Wei et al. 2007). In our previous study, Blankenship et al. (2016) reported within-frequency 2 kHz pure-tone gap detection thresholds that ranged from 5 to 70 ms (Mean = 23.2 ms, SD = 16.1). Similarly, Wei et al. (2007) reported an approximate mean GDT of 30 ms with thresholds ranging from 4 to 128 ms using broadband white noise. An even larger range of GDTs was reported by Tyler et al. (1989), where CI recipients (n = 63) displayed GDTs that ranged from 7.5 to 300 ms (Mean = 34.5 ms). They also reported that CI recipients with GDTs > 40 ms displayed poorer speech and environmental noise identification abilities compared to CI recipients with GDTs < 40 ms. Similarly, Muchnik et al. (1994) reported mean narrowband noise GDTs of 12.2 ms (SD = 18.7) in CI recipients with open-set speech recognition and a mean of 41.0 ms (SD = 34.3) in participants without significant open-set speech recognition.

The slightly higher mean GDTs reported in the literature could be due to several factors including the complexity of the signal, overall stimulus duration, and inclusion of pre-lingually deafened CI recipients. Blankenship et al. (2016) only included post-lingually deafened participants, however, the overall stimulus duration was fixed at 17 ms compared to the variable duration of 500 to 700 ms in the current study. Wei et al. (2007) and Tyler et al. (1989) both included pre-lingually deafened CI recipients which could explain the larger range of values and the stimuli were more complex (broadband and narrowband noise) potentially contributing the higher mean GDTs. Lastly, although Muchnik et al. (1994) only included post-lingually deafened individuals, 6 participants displayed no significant open-set speech recognition which likely contributed to the higher mean GDT. Overall, our NH and CI GDT results are consistent with the literature and further supports the variability seen in CI recipients.

### Behavioral Speech Perception

Normal hearing participants performed near ceiling level with CI recipients performing significantly poorer on all speech perception measures. On the CNC and AzBio, NH participants displayed mean scores that were > 99%. CI recipients that displayed mean scores that ranged from 66 to 89% correct, which is on average 11 to 34% poorer compared to NH participants. On the BKB-SIN, CI recipients required an 8.9 dB SNR increase compared to NH participants to understand 50% of the target words in the sentence.

NH group mean results are consistent with those reported in the literature. Saunders et al. (2018) reported mean CNC word scores of 95.4% in adult normal hearing participants. More recently Holder et al. (2018) conducted a large scale study of speech recognition in noise in NH adults (n = 18, Range = 20 to 79 yrs.). Participants performed near ceiling level with mean AzBio sentence recognition in noise (+10 dB) scores of 99% and BKB-SIN SNR-50 was −1.3 dB, consistent with the current study. Blankenship et al. (2016) displayed a slightly better SNR-50 of −4.2 dB for a group of young NH adults. Lastly, the normative SNR-50 value reported in the BKB-SIN test manual for normal hearing adults is −2.5 dB, similar to the current study (Etymotic 2005).

Mean speech perception scores in the CI group are also similar to those reported in previous studies. In a group of 114 post-lingually deafened CI recipients, Holden et al. (2013) reported mean CNC word scores of 61.5% (Range = 2.9 to 89.3%) at 2 years post initial activation. Similarly, Blankenship et al. (2016) reported a mean CNC word recognition score of 64.9%, CNC phoneme recognition score of 79.1%, and AzBio sentence recognition in quiet of 75.5% in a small group of post-lingually deafened CI users (n = 17 ears). Gifford et al. (2008) reported mean speech recognition scores for post-lingually deafened unilateral and bilateral adult CI users (N = 156) that included CNC word recognition (Unilateral = 55.7%, Bilateral = 65.3%), AzBio sentence recognition (Unilateral = 72.1%, Bilateral = 81.2%), BKB-SIN SNR-50 (Unilateral = 11.4 dB, Bilateral = 9.8 dB). Lastly, Donaldson et al. (2009) reported a mean BKB-SIN SNR-50 value of 11.9 dB in a group of mostly post-lingually deafened CI users. Slight differences observed in speech perception scores across studies could be due to stimulus presentation levels, age of the participants, and due to the number of unilateral and bilateral CI recipients in the study.

### Within-Frequency Electrophysiological Gap Detection

CAEPs evoked by gaps in within-frequency stimuli were examined in NH and CI participants using four gap durations that ranged from the reference condition (no gap) to a supra-threshold gap duration condition (behavioral GDT x 3). CAEP waveforms were obtained separately for both the pre- and post-gap marker. As anticipated, P1, N1, P2 were present and easily identified for the pre-gap marker for NH and CI participants for all conditions. However, compared to NH participants, CI recipients displayed smaller amplitude, and increased latency for all CAEP components. Post-gap CAEP responses were the focus of the study and are discussed in further detail below.

In NH participants, the reference condition did not elicit a post-gap CAEP response for any participant (n = 0), sub-threshold and threshold conditions showed an equal number of CAEP responses (n = 13), and all participants displayed a post-gap CAEP response to the supra-threshold condition (n = 15). Since all NH participants had a behavioral GDT of 2 ms, the gap durations for the CAEP conditions from sub-to supra-threshold were 1, 2, and 6 ms, respectively. Therefore, it is not unexpected that most NH participants would display a CAEP at the sub-threshold condition since it was only 1 ms shorter than their behavioral GDT. For NH participants, behavioral GDTs and CAEP responses were within 1 to 4 ms (Mean = 1.2 ms). Additionally, NH participants displayed a trend of increased N1-P2 amplitude for longer gap durations. For example, as the silent gap duration increased, the N1-P2 amplitude systematically increased (sub-threshold = 1.2 µV, threshold = 1.5 µV, supra-threshold = 1.8 µV).

Similarly, in CI recipients, the reference condition did not elicit a post-gap CAEP response for any participant (n = 0), sub-threshold and threshold conditions showed an equal number of present responses (n = 13), and almost all participants displayed a CAEP response to the supra-threshold condition (n = 14). There were two participants (Sci44-Left and Sci19-Right) that did not have a CAEP present for the sub-threshold condition (1 ms) or threshold condition (2 ms) but a CAEP was observed for the supra-threshold condition (6 ms). In contrast, Sci39-Right displayed a present sub- and threshold condition (1 and 2 ms respectively) but had an absent CAEP to the supra-threshold gap duration (6 ms). Furthermore, three CI recipients had elevated behavioral GDTs (Sci36-Right = 26.7 ms, Sci43-Left = 41.7 ms, and Sci36-Left = 51.6 ms) but had present sub-threshold CAEPs at 9, 14, and 17 ms, respectively. Collectively for CI recipients, behavioral GDT and CAEP responses were within 1 to 35 ms, with a mean difference of 7 ms.

Upon comparison of NH and CI recipient post-gap CAEP waveforms, CI recipients displayed poorer overall waveform morphology, smaller N1 amplitude, larger P2 amplitude, and an increased P1 latency compared to NH participants. Additionally, CI recipients did not display a similar trend of increased N1-P2 amplitude with longer silent gap durations (sub-threshold = 1.2 µV, threshold = 1.0 µV, supra-threshold = 1.2 µV) that was observed in the NH participants. Differences in CAEP amplitude and latency can be further explained by differences in neural density, adaptation, and recovery. The CAEP response is elicited by groups of neurons within the cortex that are limited in their ability to respond to acoustic stimuli by adaptation and refractory periods. For within-frequency gap detection, the neurons initially respond to the onset of the stimuli (pre-gap marker) and then are activated again by the onset of the post-gap marker. Due to the relatively short duration between the onset of the pre-gap marker and the onset of the post-gap marker in this study, the neuron is not able to fully recover. Stimuli that contain longer gap durations, thus increasing the time between the onset of the pre- and post-gap marker, would allow a more complete recovery of the neurons. Animal studies that examined cortical temporal acuity have demonstrated similar temporal mechanisms that are involved in neural recovery and gap detection (Kirby and Middlebrooks 2010). These results have also been replicated in NH adults where longer gap durations result in larger CAEP amplitudes (Lister et al. 2007; Lister et al. 2011; Palmer and Musiek 2013). With longer gap durations, full or near full recovery from adaptation is possible, thus a greater number of neurons can fire collectively resulting in increased CAEP amplitude. This supports our finding of increased CAEP amplitude with longer gap durations in NH participants. However, CI recipients did not display a similar systematic change in CAEP amplitude with longer gap duration which might be due to a reduced number of neurons that are able to respond and encode gaps due to auditory deprivation and or perhaps cortical neurons in CI recipients have increased refractory periods and are not able to fire as quickly after the pre-marker stimuli. Additionally, P1 latency was prolonged in CI recipients (Mean = 96.5 ms) compared to NH participants (Mean = 57.6 ms). P1 is a reflection of the cumulative synaptic delay from the peripheral to central auditory system (Eggermont et al. 1997; Steinschneider et al. 1994) and has been associated with auditory inhibition and sensory gating (Huotilainen et al. 1998; Thoma et al. 2003; Waldo et al. 1992). Increased P1 latency in CI recipients may suggest less efficient transmission of the auditory signal to the auditory cortex and slower neural processing of acoustic features within the stimulus.

The largest effect on CAEP amplitude was due to age with older participants displaying smaller N1-P2 amplitudes (Mean = 1.183 µV) compared to younger participants (Mean = 1.505 µV). Our findings are consistent with previous animal studies that have reported that older animals have up to 50% less cortical neurons that can encode gap and demonstrated slower recovery after gap detection (Walton et al. 1998). The combination of reduced neural processing of gaps and reduced overall number of neurons in the auditory cortex of older adults (Brody 1955) could be responsible for the smaller CAEP amplitude. In human studies, Harris et al. (2012) reported that the N1, P2, and N2 amplitudes were robustly affected by age, with younger adults displaying significantly larger amplitudes than older adults with normal hearing. In contrast, Lister et al. (2011) reported that older adults had larger P1 amplitude, increased P2 latency and broader CAEP peaks compared to young normal hearing adults. In conclusion, age related changes in the CAEP response have been reported in the literature with variable effects on component peak amplitude and latency.

### Within-Frequency Canonical Correlation Analysis

Multivariate canonical correlation analysis revealed a significant relationship between behavioral measures of speech perception and temporal processing and the supra-threshold and threshold CAEP condition separately. For the threshold CAEP condition, 86% and 76% overlapping variance was found with speech perception and behavioral GDTs. Smaller GDTs resulted in a reduced N1-P2 amplitude and poorer speech perception performance result in larger P1 amplitude and increased P1, N1, and P2 latency. For the supra-threshold CAEP condition, 63% and 52% overlapping variance was found with speech perception and behavioral GDTs. Better speech performance in noise results in a smaller P1 amplitude and decreased N1 latency with a minimal effect on N1-P2 amplitude, P1 and P2 latency. Larger within-frequency GDTs are related to increased P1 and N1-P2 amplitude, decreased P1 and N1 latency and increased P2 latency.

A few other studies have investigated the relationship between CAEPs to gaps and speech perception (He et al. 2013; He et al. 2015; Michalewski et al. 2005), however, they did not conduct canonical correlations but instead reported bivariate correlations or descriptive summaries. Michalewski et al. (2005) reported that individuals with auditory neuropathy and elevated behavioral GDTs had poorer speech understanding (sentences in quiet), however, no statistical analysis was completed. In pediatric CI recipients with auditory neuropathy, He et al. (2013) reported a significant negative correlation between electrophysiological GDTs (800 ms biphasic electric pulses with silent gaps) and open-set word recognition. Furthermore, in CI candidates with auditory neuropathy, He et al. (2015) reported a significant negative correlation between electrophysiological GDTs and open-set word recognition. However, it is important to note that the CAEP values used in the correlation analyses were electrophysiological GDT (ms) and not P1-N1-P1 peak amplitude and latency values.

### Within-Frequency Bivariate Correlation Analysis

In the current study, non-parametric spearman rank correlations revealed significant negative relationship for AzBio sentences in noise (r = −0.43, *p* = 0.010) and a significant positive correlation for SNR-50 (r = 0.43, *p* = 0.008) with behavioral within-frequency GDTs even after adjusting for multiple comparisons. These results are consistent with previous studies showing a correlation between temporal processing and speech perception in CI recipients (Blankenship et

al. 2016; Busby and Clark 1999; Muchnik et al. 1994). The significant correlation in the current study appears to be driven by three data points (Sci36-Left = 51.7 ms, Sci36-Right = 26.7 ms, Sci43-Left = 41.7 ms). Examining demographic and device data in Table 1, Sci36 is the oldest participant in the study with the longest duration of auditory deprivation (51 yrs.). Sci43 is also an older participant (60 yrs.) but only has 23 years of auditory deprivation. The standard strategy, rate, and maxima were used for both participants (ACE, 900 Hz, 8), however, it would be interesting to determine if the pulse width was widened for this participant. With regard to speech perception, Sci43 displayed the poorest performance in the CNC-Word (43%) and Phoneme (62%) and Sci36 displayed relatively high performance on the CNC-Word (Left = 80%, Right = 74%) and Phoneme (Left = 93%, Right = 86%). On the AzBio sentences in quiet, their scores ranged from 82% to 89%. However, on the AzBio sentences in noise and the BKB-SIN, these two participants were among the poorest performers with AzBio-Noise scores ranging from 40 to 56% and SNR-50 scores of 10.8 to 12.8 dB. Although group analyses are not feasible due to small sample sizes of individuals with good (n = 13) and poor within-frequency GDT (n = 3), correlation analysis supports a significant relationship between larger GDTs and poorer speech perception in noise (AzBio-Noise, SNR-50). Result indicate that adequate speech understanding in quiet may be achieve with larger GDTs, however, smaller GDTs (i.e., better temporal processing), result in better performance on speech-in-noise tasks.

### Implications and Limitations

Consistent with previous publications, our results indicate the CAEP to gaps in pure-tones could be used to as an objective measure of temporal resolution (Lister et al. 2007; Palmer and Musiek 2013, 2014). NH participants display CAEP responses to gap durations that were within 4 ms of their behavioral threshold. CI recipients displayed more variability with 12 participants displaying CAEP responses to gap durations that were within 4 ms and 3 participants with CAEP responses to gap durations that were 24 to 35 ms smaller than their behavioral GDTs. Future studies are needed to evaluate gap evoked CAEPs in CI recipients immediately surrounding threshold with predetermined step sizes (Ex: 5 to 10 ms). For clinical applications it is important to know how closely behavioral and electrophysiological GDTs agree and the systematic change in CAEP response amplitude and latency.

The main limitation in this study is the ceiling effect observed on the within-frequency behavioral gap detection task. All NH participants (n = 15 ears) and most of the CI recipients (n = 12 ears) displayed a behavioral GDT of 2 ms, the smallest possible value on the task. Future studies should include smaller gap durations, or decrease the pre- and post-gap marker duration, thereby increasing the difficulty of the task and increase observed GDTs. Ceiling affects were also observed for NH participants on speech perception tasks. More difficult speech perception tasks such as the TIMIT sentences (commissioned by Texas Instruments and Massachusetts Institute of Technology), could be used to better reflect real life listening situations for both NH and CI recipients (Fu et al. 2002; King et al. 2012; Loizou et al. 2000; Shannon et al. 2002). In addition, for presentation in noise, the SNR could be varied to find optimal balance between SNR levels and performance for both NH and CI recipients to avoid both ceiling and floor effects. Alternatively, the Digits-in-Noise Test, which displays high repeatability and is quick and easy to administer, could be used to measure speech-in-noise (Cullington and Agyemang-Prempeh 2017; Zhang et al. 2019). The lack of variability in the within-frequency and behavioral speech perception tasks, partially undermined correlation analysis in the present study.

## CONCLUSIONS

CAEP to gaps in tones can be used to evaluate within-frequency temporal processing in NH and CI recipients. NH participants show the anticipated trend of reduced N1-P2 amplitude as CAEP gap durations decrease. CI recipients did not display this same gap duration trend but instead displayed reduced N1 amplitude and increased P2 amplitude and P1 latency compared to NH participants. There was a significant effect of age at test, with older adults displaying smaller N1-P2 amplitude than younger adults. Canonical correlations reported a strong relationship between CAEP amplitude and latency values for the threshold condition and behavioral measures of speech perception and temporal processing. Lastly, there was a significant relationship between poorer temporal processing (larger GDTs) and poorer performance on the AzBio sentences in noise and the SNR-50 with no significant correlations for words or sentences in quiet. Individuals with poorer temporal processing are likely to have adequate speech perception in quiet but worse speech understanding in noise. Additional studies need to be completed with more difficult behavioral GDT stimuli and speech perception materials and smaller CAEP gap durations near threshold that are consistent across participants.

## Data Availability

The data can be made available upon request.

## List of Abbreviations

AzBio: Arizona Biomedical Sentence Test
BKB-SIN: Bamford-Kowal-Bench Speech-in-Noise Test
CAEP: Cortical Auditory Evoked Potential
CI: Cochlear Implant
CNC: Consonant-Nucleus-Consonant Word Test
EEG: Electroencephalographic
GDT: Gap Detection Threshold
ICA: Independent Component Analysis
MCL: Most Comfortable Level
NH: Normal Hearing
SL: Sensation Level
SNR: Signal-to-Noise Ratio

## Acknowledgements

The authors thank all participants in this research. This research was supported by the National Institute Health, National Institute on Deafness and Other Communication Disorders Grant R15 DC011004 (F.Z.) and the University of Cincinnati University Research Council (C.M.B). Portions of this article were presented at the Association for Research in Otolaryngology 37^th^ and 42^nd^ Annual MidWinter Meeting in 2014 and 2019. This manuscript represents a portion of the doctoral dissertation of the first author under guidance of the co-authors. The content is solely the responsibility of the authors and does not necessarily represent the official views of the National Institutes of Health or the University of Cincinnati.

## Author Contributions

C.M.B designed and performed the experiments, analyzed the data and wrote the paper. J.M provided statistical analysis. F.Z. designed the experiments and provided interpretive analysis and critical revision to the paper.

